# Phenome-wide Mendelian randomization study of plasma triglycerides and 2,600 disease traits

**DOI:** 10.1101/2022.07.21.22277900

**Authors:** Joshua K. Park, Shantanu Bafna, Iain S. Forrest, Áine Duffy, Carla Marquez-Luna, Ben O. Petrazzini, Ha My Vy, Daniel M. Jordan, Marie Verbanck, Jagat Narula, Robert S. Rosenson, Ghislain Rocheleau, Ron Do

**Affiliations:** Charles Bronfman Institute for Personalized Medicine, Icahn School of Medicine at Mount Sinai, New York, NY, USA; Department of Genetics and Genomic Sciences, Icahn School of Medicine at Mount Sinai, New York, NY, USA; Medical Scientist Training Program, Icahn School of Medicine at Mount Sinai, New York, NY, USA; Université Paris Cité, UR 7537 BioSTM, Paris, France; Department of Medicine, Icahn School of Medicine at Mount Sinai, New York, NY; Cardiovascular Imaging Program, Zena and Michael A. Wiener Cardiovascular Institute, Mount Sinai Heart, Icahn School of Medicine at Mount Sinai, New York, NY; Metabolism and Lipids Unit, Zena and Michael A. Wiener Cardiovascular Institute, Mount Sinai Heart, Icahn School of Medicine at Mount Sinai, New York, NY

**Keywords:** Cardiovascular disease, Coronary artery disease, Dyslipidemia, FinnGen, Hypertriglyceridemia, Lipids, Mendelian randomization, Phenome-wide MR, Triglyceride-rich lipoprotein, Triglycerides, UK Biobank

## Abstract

**Background:** Causality between plasma triglyceride (TG) levels and atherosclerotic cardiovascular disease (ASCVD) risk remains controversial despite more than four decades of study and two recent landmark trials, STRENGTH and REDUCE-IT. Further unclear is the association between TG levels and non-atherosclerotic diseases across organ systems.

**Methods:** Here, we conducted a phenome-wide, two-sample Mendelian randomization (MR) analysis using inverse-variance weighted (IVW) regression to systematically infer the causal effects of plasma TG levels on 2,600 disease traits in the European ancestry population of UK Biobank. For replication, we externally tested 221 nominally significant associations (*p* < 0.05) in an independent cohort from FinnGen. To account for potential horizontal pleiotropy and the influence of invalid instrumental variables, we performed sensitivity analyses using MR-Egger regression, weighted median estimator, and MR-PRESSO. Finally, we used multivariable MR controlling for correlated lipid fractions to distinguish the independent effect of plasma TG levels.

**Results:** Our results identified 7 disease traits reaching Bonferroni-corrected significance in both the discovery (*p* < 1.92 × 10^-5^) and replication analyses (*p* < 2.26 × 10^-4^), supporting a causal relationship between plasma TG levels and ASCVDs, including coronary artery disease (OR 1.33, 95% CI 1.24-1.43, *p* = 2.47 × 10^-13^). We also identified 12 disease traits that were Bonferroni-significant in the discovery or replication analysis and at least nominally significant in the other analysis (*p* < 0.05), identifying plasma TG levels as a novel risk factor for 9 non-ASCVD diseases, including uterine leiomyoma (OR 1.19, 95% CI 1.10-1.29, *p* = 1.17 × 10^-5^).

**Conclusions:** Taking a phenome-wide, two-sample MR approach, we identified causal associations between plasma TG levels and 19 disease traits across organ systems. Our findings suggest unrealized drug repurposing opportunities or adverse effects related to approved and emerging TG-lowering agents as well as mechanistic insights for future study.

## Introduction

Atherosclerotic cardiovascular disease (ASCVD) remains the leading cause of death worldwide despite the effectiveness of statin therapy in reducing low-density lipoprotein cholesterol (LDL-C) levels (Baigent et al., 2005) (Roth et al., 2020). Additional therapeutic targets and adjunctive treatments are needed to address the burden arising from residual risk (Rosenson & Goonewardena, 2021). Triglycerides (TG) play vital roles in physiology, ranging from energy storage and mobilization to inflammation, thrombosis, and hormone-like signaling (Zewinger et al., 2020) (Norata et al., 2007). However, a causal relationship between TGs and ASCVDs remains controversial (Miller et al., 2011) (Albrink & Man, 1959), recently culminating in the conflicting reports of two double-blinded randomized controlled trials (RCT), STRENGTH and REDUCE-IT (Nicholls et al., 2020) (Bhatt et al., 2019) (Doi et al., 2021). Nevertheless, drug development for reducing TG-rich lipoproteins (TRL) is an active area of research and several targets have now been validated, including angiopoietin-like 3 (ANGPTL3) (Graham et al., 2017) (Dewey et al., 2017) (Musunuru et al., 2010) (Gaudet et al., 2017), angiopoietin-like 4 (ANGPTL4) (Dewey et al., 2016), and apolipoprotein C-III (APOC3) (Gaudet et al., 2015) (Crosby et al., 2014). Clinical trials are currently evaluating these targets for dyslipidemias (Arrowhead, 2021b) (Arrowhead, 2021a).

Whether TGs are causal risk factors or simply associative biomarkers remains uncertain not only for ASCVDs but also other diseases of different organ systems. Understanding the causal effects of TGs across a broader range of human diseases could have significant implications for drug repurposing. TG-lowering agents, such as fibrates (Frick et al., 1987) (Group, 2010) and omega-3 fatty acids (Bhatt et al., 2019) have already been approved; however, not knowing which diseases are causally affected by TGs precludes their use for indications other than hypertriglycidemia. Understanding the protective effects of TGs could also have implications for drug safety. With the recent approval of icosapent ethyl (Bhatt et al., 2019) (Gaba et al., 2022) and the ongoing development of other TG-lowering agents (Shaik & Rosenson, 2021), such as ANGPTL3 (Graham et al., 2017) (Dewey et al., 2017), ANGPTL4, and APOC3 inhibitors (Gaudet et al., 2015), long-term safety becomes a matter of concern. Post-market surveillance data are limited; therefore, identifying diseases with negative causal links to TGs could suggest adverse side effects, whereas protective causal links to TGs could suggest therapeutic avenues and indices informative for drug development. Further, this could inform polypharmacy and drug titration in clinical practice.

Several methodological challenges have prohibited causal conclusions about TGs across human diseases. First, TGs are correlated with established ASCVD risk factors, such as obesity and insulin resistance (Eckel et al., 2005). They also correlate with LDL particles or apolipoprotein B (apoB) concentration (Cromwell et al., 2007) and inversely correlate with high-density lipoprotein cholesterol (HDL-C) (Phillips & Smith, 1991). Conventional observational studies have thus been limited in drawing causal inferences due to potential confounding. Secondly, available TG-lowering agents have pleiotropic effects on several major lipid fractions, including VLDL-C, LDL-C, HDL-C, and apolipoproteins, including APOC3 (Group, 2010) (Keech et al., 2005) (Frick et al., 1987). Thus, costly RCTs have had limited power to disentangle the effects of lowering TG specifically (Triglyceride Coronary Disease Genetics et al., 2010) (Goldberg et al., 2011) (Bhatt et al., 2019). Finally, Mendelian randomization (MR) studies have typically focused on individual diseases selected *a priori* (Davey Smith & Hemani, 2014), narrowing the scope of causal estimates to ASCVDs while overlooking non-ASCVD diseases (Harrison et al., 2018) (Smith et al., 2014) (Allara et al., 2019).

Phenome-wide MR is a high-throughput extension of MR that, under specific assumptions, estimates the causal effects of an exposure on multiple outcomes simultaneously. As in conventional MR, this method uses genetic variants as instrumental variables (IV) to proxy modifiable exposures while minimizing confounding (Davey Smith & Ebrahim, 2003). A distinction, however, is that phenome-wide MR enables comprehensive scans of the phenotypic spectrum, limiting bias from prior assumptions and facilitating the discovery of unforeseen causal relationships. This has recently become feasible with the maturation of large-scale biobanks providing extensive genetic and phenotypic data, such as the UK Biobank (UKB) (Bycroft et al., 2018) and FinnGen project (FinnGen, 2020).

Here, we use phenome-wide MR to systematically estimate the causal effects of plasma TGs on 2,600 disease outcomes in UKB, followed by replication testing in FinnGen. We then apply multiple MR methods for sensitivity analyses and multivariable MR to control for plasma LDL-C and HDL-C level.

## Methods

### Study design and data sources

We followed the guidelines published by Burgess et al. (Burgess et al., 2020) and the Strengthening the Reporting of Observational Studies in Epidemiology (STROBE) guidelines (Skrivankova et al., 2021). Accordingly, we note that MR analyses rely on three important instrumental variable assumptions: (1) the genetic variant is directly associated with the exposure; (2) the genetic variant is unrelated to confounders between the exposure and outcome; and (3) the genetic variant has no effect on the outcome other than through the exposure (Davey Smith & Ebrahim, 2003). This study uses three non-overlapping genome-wide association studies (GWAS). A schematic figure summarizes the study design (Fig 1).

**Fig 1.**
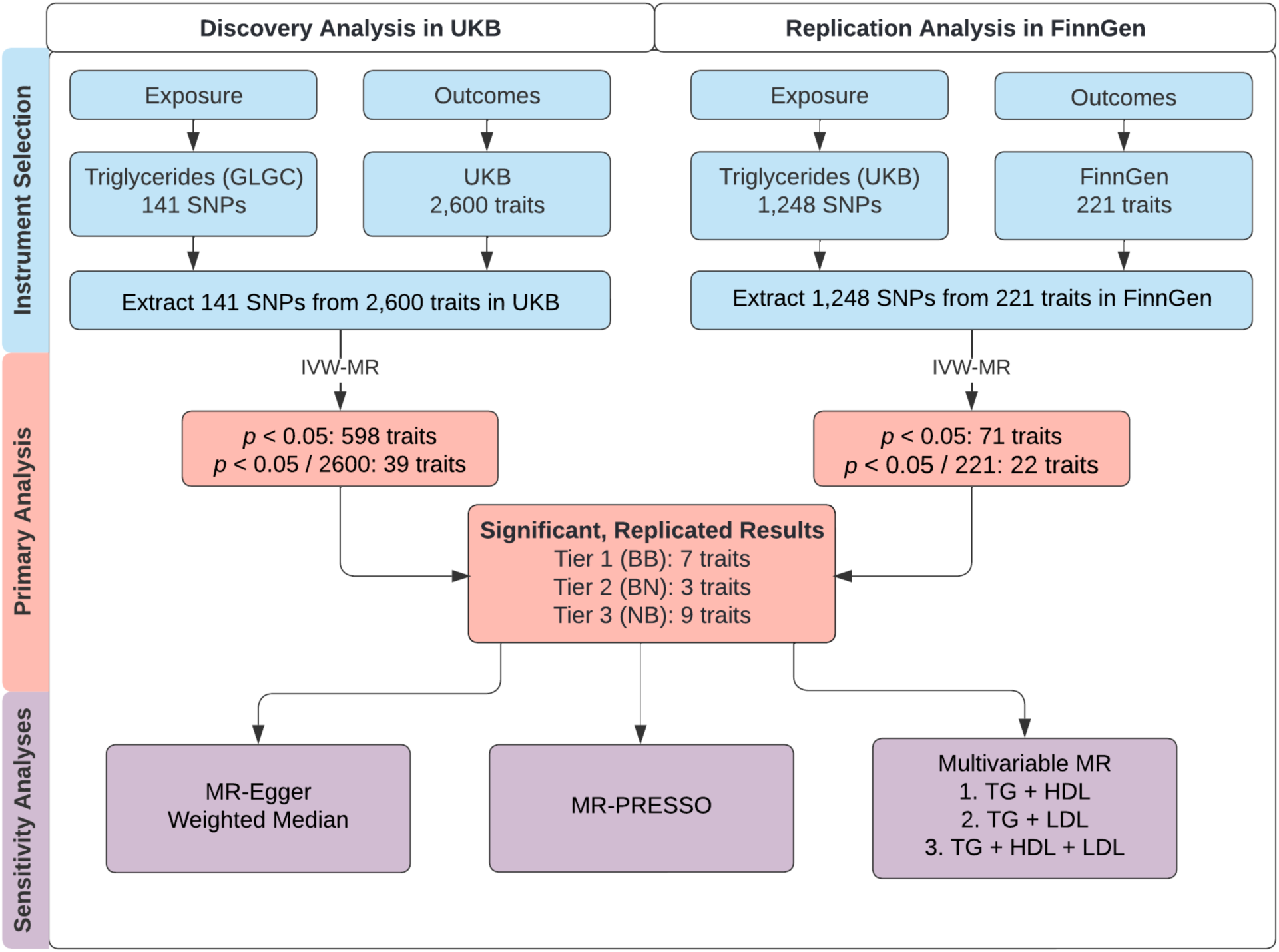
Study overview. A schematic summarizing the study design. GLGC, Global Lipids Genetics Consortium; HDL, high-density lipoprotein; IVW, inverse-variance weighted; LDL, low-density lipoprotein; MR, Mendelian randomization; MR-PRESSO, Mendelian Randomization Pleiotropy RESidual Sum and Outlier; SNP, single-nucleotide polymorphism; TG, triglyceride; UKB, UK Biobank. Tier 1 (BB): At least Bonferroni-significant in both the discovery and replication analyses. Tier 2 (BN): At least Bonferroni-significant in the discovery analysis and at least nominally significant in the replication analysis. Tier 3 (NB): At least nominally significant in the discovery analysis and at least Bonferroni-significant in the replication analysis.

For primary and sensitivity analyses, we conducted two-sample MR taking summary statistics for genetic associations with plasma TGs from one dataset (Global Lipids Genetics Consortium, GLGC) (Willer et al., 2013) and summary statistics for genetic associations with outcomes from a second, independent dataset (UKB). The CARDIoGRAMplusC4D Metabochip study by GLGC involved 63,746 cases and 130,681 controls (Deloukas et al., 2013). UKB is a longitudinal, population-based cohort study with genetic and phenotypic data on over 500,000 participants aged 40-69 years at recruitment from across the United Kingdom during 2006-2010 (Sudlow et al., 2015). The sociodemographic and health-related characteristics of UKB participants have been described elsewhere (Fry et al., 2017).

For replication analysis, we again performed two-sample MR but used summary statistics for TG from UKB and summary statistics for outcomes from FinnGen (Release 4; 176,899 samples; 169,962,023 variants). FinnGen is a large public-private partnership started in August 2017 aiming to collect and analyze genomic and phenotypic data from 500,000 Finnish biobank participants (FinnGen, 2020) (Kurki et al., 2022). For both primary and replication analysis, there is likely to be minimal sample overlap in the two-sample MR, which can cause weak-instrument bias and inflated type 1 error (Burgess et al., 2016).

### Genetic instruments

For primary and secondary analyses, we identified 3,086 single nucleotide polymorphisms (SNP) associated with plasma TG levels in GLGC at a genome-wide significance threshold of *p* < 5 × 10^-8^. A total of 141 independent SNPs were then selected at a linkage disequilibrium (LD) threshold of r^2^ < 0.05 using the 1000 Genomes LD European panel as the reference population (Auton et al., 2015). To select independent SNPs, we used the PLINK software’s LD clumping command (Purcell et al., 2007) with the following options: -- clump-p1 0.00000005 --clump-p2 0.0005 --clump-r2 0.05 --clump-kb 250.

For replication analysis, we identified 1,388 SNPs associated with plasma TG levels in UKB after restricting for genome-wide significance (*p* < 5 × 10^-8^) and LD-clumping. The SNPs were selected at an LD threshold of r^2^ < 0.05 using the 1000 Genomes LD European panel as the reference population. LD clumping was performed by PLINK using the same command as above. We then restricted to 1,248 SNPs for genetic instrumentation based on whether the SNP was also available in the FinnGen dataset.

### Disease outcomes

As a phenome-wide investigation, this study examined many binary disease outcomes. For outcomes of primary and sensitivity analyses, we used genome-wide association summary statistics from the European ancestry subset of UKB. The focus on European ancestry was needed to reduce heterogeneity and maximize statistical power. Pan-UKB performed 16,000 genome-wide association studies in ∼500,000 UKB participants of various ancestries using genetic and phenotypic data (PanUKBTeam, 2020). A total of 7,221 total phenotypes were categorized as “biomarker”, “continuous”, “categorical”, “ICD-10 code”, “phecode”, or “prescription” (PanUKBTeam, 2020). We filtered for outcomes to retain categorical, ICD-10, and phecode types; non-null heritability in European ancestry as estimated by Pan-UKB; and relevance to disease, excluding medications. This yielded 2,600 traits for primary analysis.

For outcomes of replication analysis, we used genome-wide association summary statistics from FinnGen. To allow for compatibility between UKB and FinnGen datasets, phenotypes coded as FinnGen “endpoint IDs” were mapped to ICD-10 codes or phecodes corresponding to the coding convention used by UKB. Categorical traits in UKB could not be reliably mapped to FinnGen IDs, requiring us to omit categorical traits from replication analysis. Of 2,444 available FinnGen outcomes, we ultimately selected 221 outcomes for replication testing based on two conditions: (1) The outcome has an equivalent phenotype documented as an ICD-10 code or phecode in UKB, which we had included in the discovery analysis, and (2) the outcome was at least nominally significant (*p* < 0.05) in the discovery analysis.

### Statistical analyses

Primary discovery analysis used the inverse-variance weighted (IVW) method of two-sample MR over the more horizontal pleiotropy-robust MR-Egger regression method to maximize statistical power (Burgess et al., 2013). To proxy TG, we selected the 141 TG-associated SNPs from Pan-UKB identified above as instrumental variables in MR testing. All SNPs were harmonized to match the effect and non-effect allele at each SNP. For replication analyses, we conducted IVW tests using TG-associated SNPs from UKB as exposures and 221 traits at least nominally significant (*p* < 0.05) in the discovery analysis as outcomes from FinnGen. R (4.1.0) was used for all analyses (R Core Team, 2021).

For sensitivity analyses, we addressed the possible presence of horizontal pleiotropy by applying MR-Egger (Bowden et al., 2015), weighted median estimator (Bowden et al., 2016), and MR-PRESSO outlier tests (Verbanck et al., 2018) on Bonferroni-significant traits identified in the primary analysis above. MR-PRESSO outlier tests were performed on 16 traits with significant MR-PRESSO global test results (*p* < 0.05). To account for potential horizontal pleiotropy due to outliers, we examined the MR-PRESSO outlier test p-values from the discovery analysis, and as recommended (Burgess et al., 2020), removed outlier IVs (between 1 and 10 SNPs for each outcome) based on a corrected significance threshold (*p* < 0.05 / 141 = 3.55 × 10^-4^). IVW tests were then re-run without the outlier IVs in the discovery analysis.

Additionally, we fitted multivariable MR (MVMR) models controlling for plasma LDL-C and HDL-C levels in the discovery stage. MVMR uses genetic variants associated with multiple exposures to estimate the effect of each exposure on an outcome, thereby accounting for potentially correlated exposures (Sanderson, 2021). We extracted the 141 LD-independent TG-associated SNPs described above from the summary statistics for LDL and HDL provided by GLGC. We then performed three MVMR IVW tests against traits Bonferroni-significant in our discovery analysis with the following sets of exposures: TG and LDL; TG and HDL; or TG, HDL, and LDL.

To account for multiple testing, a conservative Bonferroni threshold for statistical significance was set to *p* < 0.05 / 2600 = 1.92 × 10^-5^ for the discovery analysis. For replication analysis, a similar conservative Bonferroni threshold was set to *p* < 0.05 / 221 = 2.26 × 10^-4^. We determined three tiers of statistical evidence based on the Bonferroni or nominal threshold (*p* < 0.05) in both the discovery and replication analyses. These three tiers are defined below:

1. Tier 1 (BB): At least Bonferroni-significant in both the discovery and replication analyses.
2. Tier 2 (BN): At least Bonferroni-significant in the discovery analysis and at least nominally significant in the replication analysis.
3. Tier 3 (NB): At least nominally significant in the discovery analysis and at least Bonferroni-significant in the replication analysis.

### Ethical approval

UK Biobank has approval from the North West Multi Centre Research Ethics Committee (MREC) as a Research Tissue Bank (RTB) (11/NW/0382), and all participants of UKB provided written informed consent. More information is available at (https://www.ukbiobank.ac.uk/learn-more-about-uk-biobank/about-us/ethics). The work described in this study was approved by UKB under application number 16218. All participants of FinnGen provided written informed consent for biobank research, based on the Finnish Biobank Act. The Coordinating Ethics Committee of the Hospital District of Helsinki and Uusimaa (HUS) approved the FinnGen study protocol Nr HUS/990/2017. More information is available at (https://www.finngen.fi/en/code_of_conduct).

### Data availability

All data generated in this study are included in the manuscript and supplementary tables.

All analyses used publicly available data (UKB, FinnGen), including previously published GWAS (GLGC) (Willer et al., 2013). Obtaining access to UKB (PanUKBTeam, 2020) and FinnGen (FinnGen, 2020) GWAS summary statistics is detailed here (https://www.finngen.fi/en/access_results) and here (https://www.ukbiobank.ac.uk/enable-your-research/apply-for-access). Please note the summary statistics for FinnGen and Pan-UKB are made publicly available.

## Results

### Genetically proxied plasma TG levels and phenome-wide disease risk

In the discovery analysis, we identified nominally significant associations (*p* < 0.05) between plasma TG levels and 598 disease traits in UKB (S1 Table). Of these, 39 disease traits were statistically significant after multiple testing correction with a conservative Bonferroni-corrected threshold (*p* < 1.92 × 10^-5^). As a positive control, plasma TG levels were positively associated with gout with an odds ratio (OR) of 1.78 for gout (95% CI 1.52-2.09, *p* = 7.41 × 10^-11^), in agreement with prior studies (Yu et al., 2021). In the replication analysis, we identified nominally significant associations (*p* < 0.05) between plasma TG levels and 71 disease traits in FinnGen (S2 Table). Of these, 22 traits were Bonferroni-significant. A summary of the 19 most statistically significant and replicated results have been organized into three predefined tiers of evidence (Fig 2). We note that the magnitude of estimates from the discovery analysis were generally greater than those from replication analysis, especially among tier 1 and tier 2 associations, potentially as a manifestation of winner’s curse bias (Göring et al., 2001).

**Fig 2.**
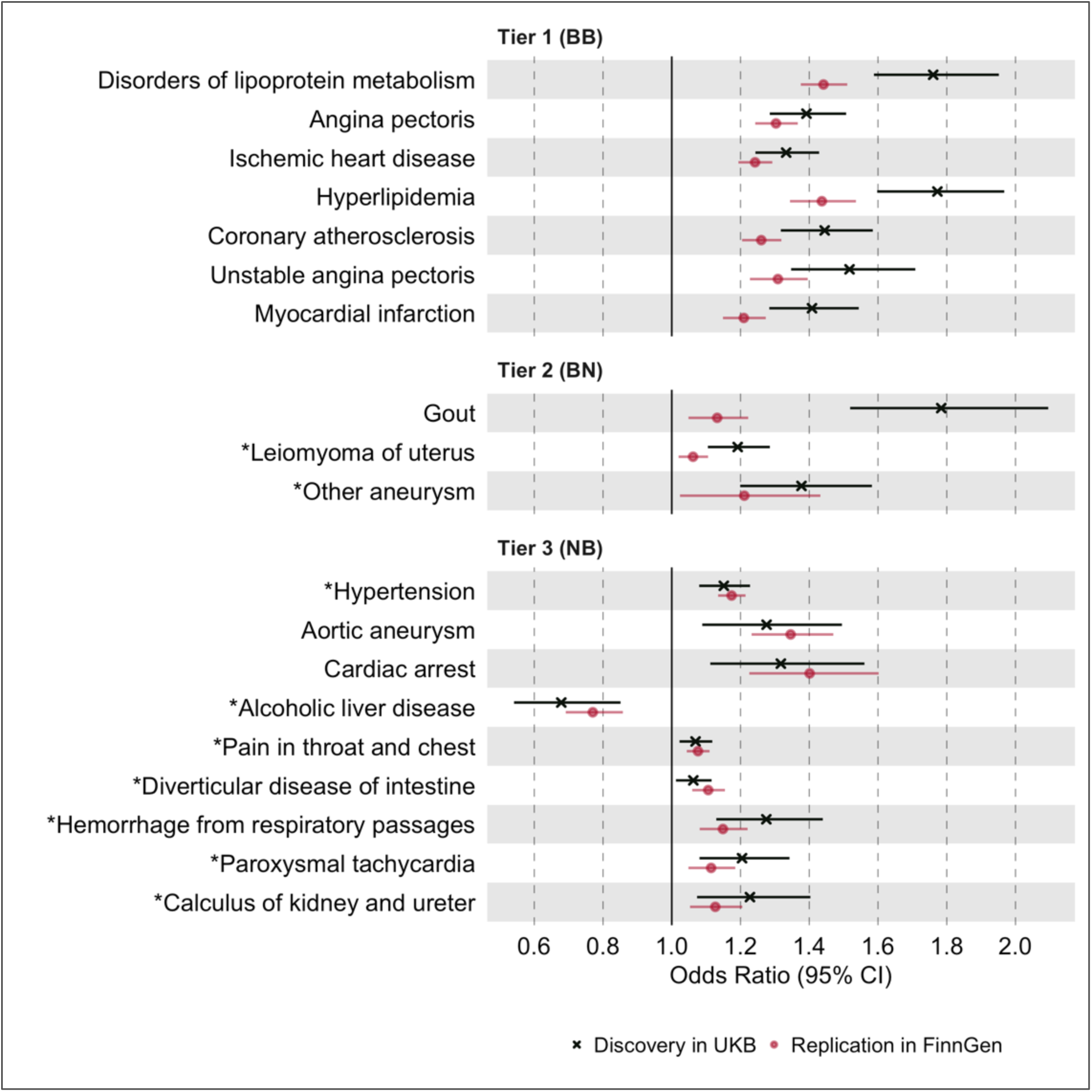
Causal estimates of genetically proxied plasma TG levels on disease risk using IVW regression in UKB and FinnGen. Causal estimates from inverse-variance weighted (IVW) regression are shown as odds ratios (OR) per 1 SD increase in plasma triglyceride (TG) levels (mmol/L). Asterisks indicate novel associations. Black indicates discovery analysis results using UKB. Red indicates replication analysis results using FinnGen. Horizontal lines represent 95% confidence intervals. Tier 1 (BB): At least Bonferroni-significant in both the discovery and replication analyses. Tier 2 (BN): At least Bonferroni-significant in the discovery analysis and at least nominally significant in the replication analysis. Tier 3 (NB): At least nominally significant in the discovery analysis and at least Bonferroni-significant in the replication analysis. Full results are provided in S1 and S2 Tables.

For tier 1 results, genetically determined plasma TG levels were positively associated with 7 disease traits in both the discovery and replication analyses. These were Bonferroni-significant in both analyses, and all were related to dyslipidemias or ASCVD. Among traits related to ASCVD, the strongest association by statistical significance was for angina pectoris in both the discovery UKB cohort (OR 1.39, 95% CI 1.29-1.51, *p* = 2.11 × 10^-13^) and the replication FinnGen cohort (OR 1.30, 95% CI 1.24-1.37, *p* = 1.25 × 10^-26^). For tier 2 results, plasma TG levels were positively associated with 3 disease traits, including non-ASCVDs: gout, uterine leiomyoma, and “other aneurysms” (phecode-442). The strongest association by significance, after gout, was for leiomyoma of the uterus in both the discovery (OR 1.19, 95% CI 1.10-1.29, *p* = 1.17 × 10^-5^) and replication cohorts (OR 1.06, 95% CI 1.02-1.11, *p* = 3.60 × 10^-3^). For tier 3 results, 9 disease traits were identified, and a greater proportion were non-ASCVD traits. The strongest association by significance was for hypertension in both the discovery (OR 1.51, 95% CI 1.08-1.23, *p* = 3.20 × 10^-5^) and replication cohorts (OR 1.17, 95% CI 1.14-1.22, *p* = 9.49 × 10^-20^).

### Sensitivity analyses using MR-PRESSO, MR-Egger, and Weighted Median methods

To account for potential horizontal pleiotropy bias due to outlier IVs, we next used the MR-PRESSO test on the 19 significant and replicated associations identified above, which were categorized into three tiers of statistical evidence. MR-PRESSO suspected outlier IVs for 16 of the 19 associations, and between 1 and 10 IVs were identified for 8 associations (S3 Table). We then re-ran IVW MR for these 8 associations after removing outlier IVs (S3 Table). Among tier 1 results, all 7 associations increased in significance after outlier removal (Fig 3). For tier 2 results, two of three associations had significant global MR-PRESSO test values but no outlier IVs were removed. For tier 3 results, seven of nine associations had significant global MR-PRESSO test values, and outlier IVs were removed for one of these associations (S3 Table), which increased in significance: hypertension (*p* = 3.20 × 10^-5^ to *p* = 2.80 × 10^-8^). Importantly, all associations maintained the same effect direction after the MR-PRESSO outlier test, in keeping with the initial discovery analysis.

**Fig 3.**
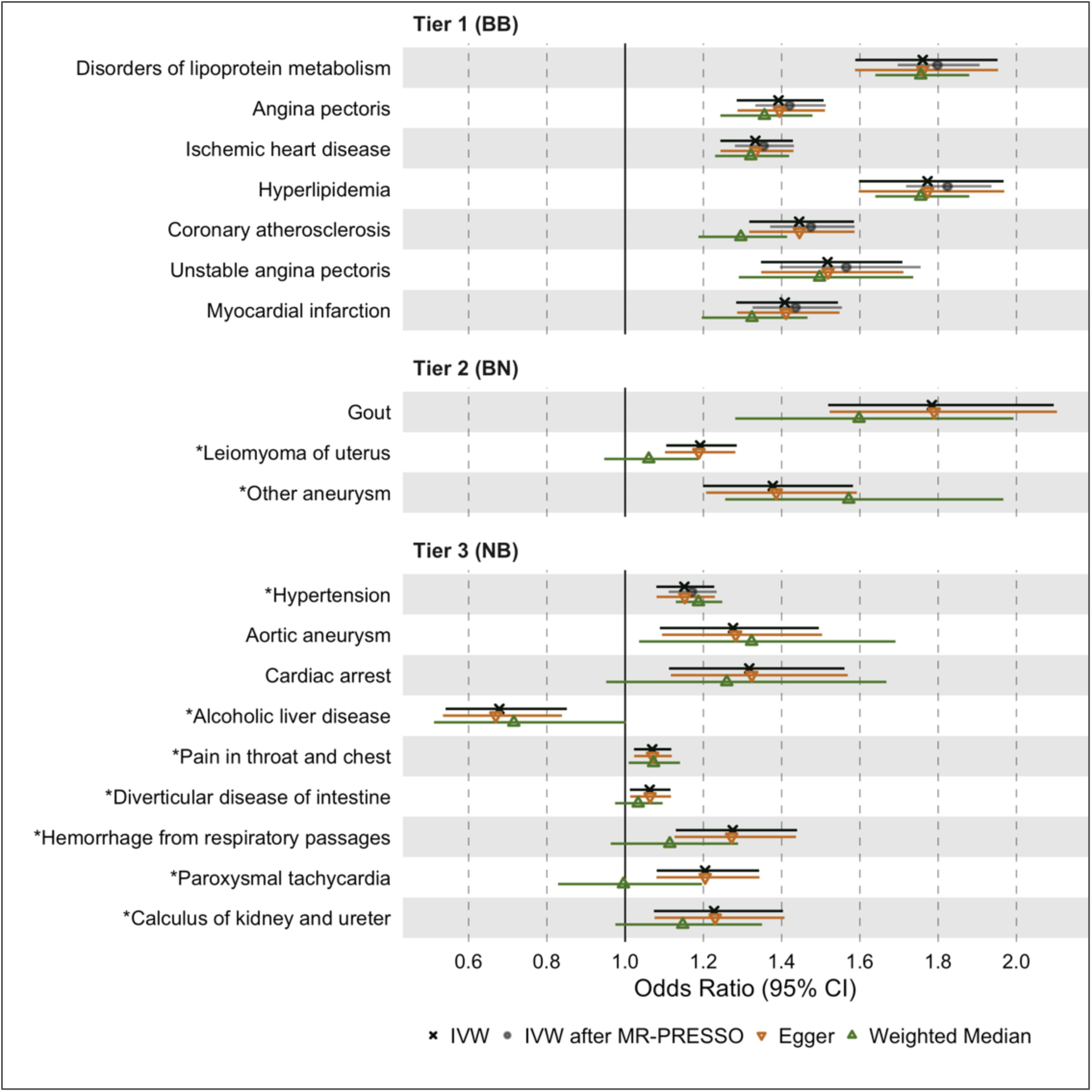
Causal estimates of genetically proxied plasma TG levels on disease risk using MR-Egger, Weighted Median, and MR-PRESSO methods in UKB. Shown are sensitivity analysis results from the discovery stage using instruments from GLGC and outcomes from UKB. Levels of statistical significance are expressed as -log_10_(*p*) values. Associations with insignificant global MR-PRESSO test results (*p* > 0.05) were not rerun and do not have data points for IVW after MR-PRESSO. Tier 1 (BB): At least Bonferroni-significant in both the discovery and replication analyses. Tier 2 (BN): At least Bonferroni-significant in the discovery analysis and at least nominally significant in the replication analysis. Tier 3 (NB): At least nominally significant in the discovery analysis and at least Bonferroni-significant in the replication analysis. Full results are found in S1 and S3 Tables.

To further account for horizontal pleiotropy, we conducted sensitivity analyses for the 19 significant and replicated traits using the MR-Egger and weighted median estimators on UKB data (Fig 3). MR-Egger results were comparable to IVW results for all 19 traits across the three tiers of evidence. However, weighted median results were less statistically significant than IVW and MR-Egger results for most traits except for disorders of lipid metabolism, hyperlipidemia, and hypertension.

### Separating the independent effects of plasma TG levels

To isolate the independent effect of plasma TG levels from those of correlated lipid fractions, we next conducted multivariable MR (MVMR) on the 19 significant and replicated associations using the IVW estimator on UKB data, adjusting for plasma HDL-C, LDL-C, or both simultaneously (Fig 4). For tier 1 associations, we observed an increase in statistical significance for disorders of lipid metabolism and hyperlipidemia after adjustment. For tier 2 results, we observed an increase in significance for gout. For tier 3 results, we observed an increase in significance for alcoholic liver disease, paroxysmal tachycardia, and calculus of kidney and ureter. Aside from these, MVMR led to a decrease in statistical significance for the remaining associations. However, all 19 associations remained in the same direction of causality despite multivariable adjustment (S4 Table).

**Fig 4.**
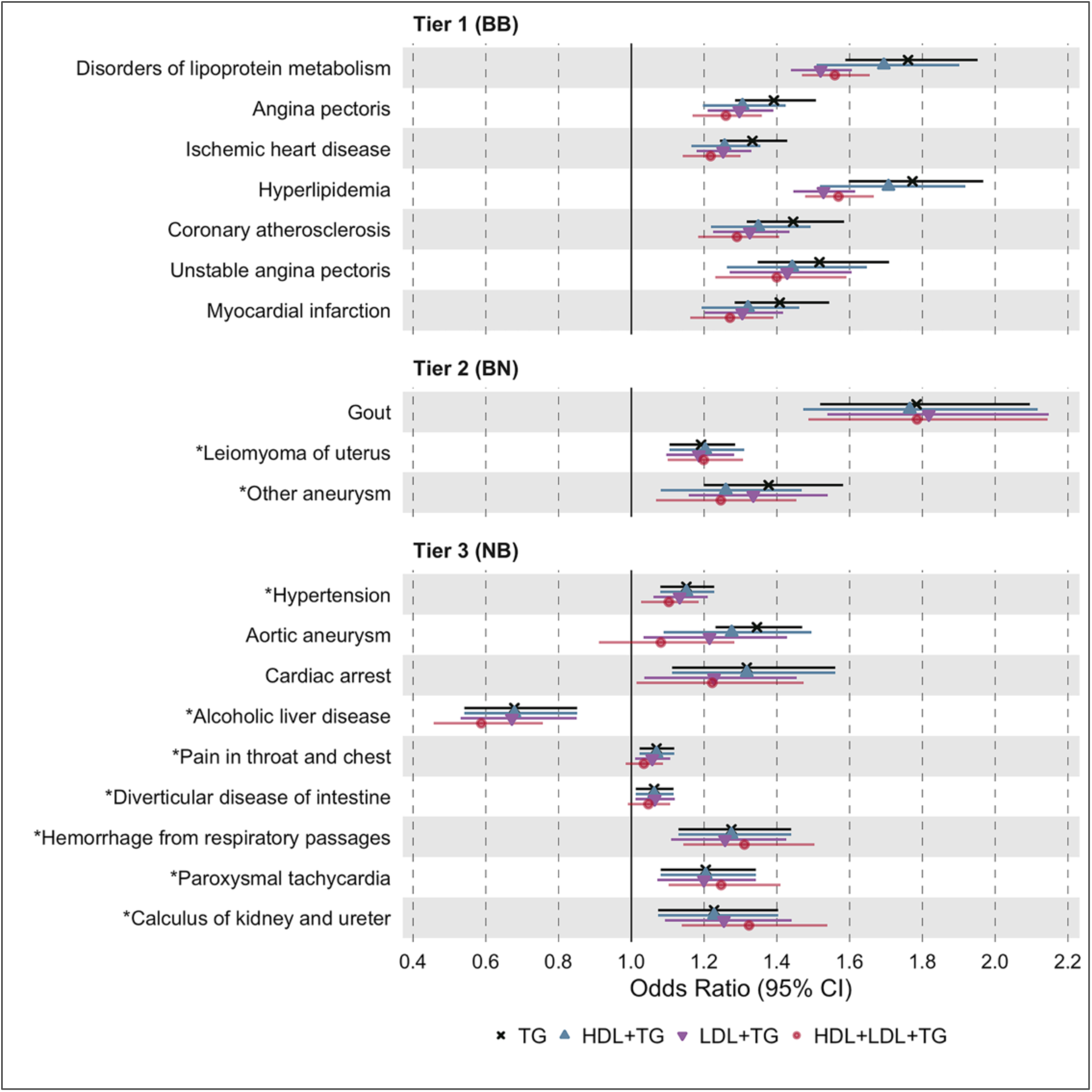
Causal estimates of genetically proxied plasma TG levels on disease risk using multivariable IVW regression controlling for plasma LDL-C and HDL-C levels in UKB. Shown are the multivariable MR results from the discovery analysis, using genetic instruments for plasma TG levels from GLGC and disease traits from UKB. Degrees of statistical significance are categorized into three tiers: Tier 1 (BB): At least Bonferroni-significant in both the discovery and replication analyses. Tier 2 (BN): At least Bonferroni-significant in the discovery analysis and at least nominally significant in the replication analysis. Tier 3 (NB): At least nominally significant in the discovery analysis and at least Bonferroni-significant in the replication analysis. Full results are provided in S4 Table.

## Discussion

We performed phenome-wide, two-sample MR to estimate the causal effects of plasma TG levels on a spectrum of human disease traits (*n* = 2,600) using a discovery cohort from UKB and a replication cohort from FinnGen. We report 7 disease traits reaching Bonferroni-corrected significance in both the discovery and replication analyses, which we categorized as tier 1 results. These traits were predominantly manifestations of ASCVD, such as ischemic heart disease, angina pectoris, and myocardial infarction. Using multivariable MR, we found that these associations remained even after controlling for LDL-C and HDL-C (Fig 4). We also identified 3 disease traits that are both Bonferroni-significant in the discovery analysis (*p* < 1.92 × 10^-5^) and at least nominally significant in the replication analysis (*p* < 0.05), categorized as tier 2 results. Lastly, we identified 9 disease traits at least nominally significant in the discovery analysis and Bonferroni-significant in replication analysis, categorized as tier 3 results. Several of these disease traits have never been reported to be causally associated with TGs, introducing new opportunities for drug repurposing and further mechanistic studies.

Our results are consistent with prior work suggesting that plasma TG levels are causally associated with ASCVD risk (Triglyceride Coronary Disease Genetics et al., 2010) (Castañer et al., 2020) (Do et al., 2013) (Dewey et al., 2016) (Holmes et al., 2015) (White et al., 2016) (Varbo et al., 2013) (Ibi et al., 2021) (Rosenson et al., 2021). We do not prove here that circulating TGs *per se* are directly atherogenic; rather, we show that plasma TG measurement, which comprises multiple classes of triglyceride-rich lipoproteins (TRL) and their remnants, captures a clinically significant mechanism that is causally associated with ASCVD (Rosenson et al., 2021). This interpretation is concordant with the emerging consensus that apolipoprotein B (apoB) particle number is a more important determinant of atherogenesis than LDL-C and that TRL particles are as important a risk factor in ASCVD as LDL particles, since they both carry a single apoB molecule (Ference et al., 2019) (Marston et al., 2021) (Richardson et al., 2020). According to this paradigm, the causal association we observe between plasma TG levels and ASCVD risk may be mediated by the concentration of apoB particles in TRLs, captured by plasma TG level measurements. If reducing TRLs proportionally reduces apoB levels (Ference et al., 2019), these results support that lowering plasma TG levels could serve as a valid therapeutic strategy to reducing ASCVD risk. Additional studies controlling for apoB are needed to test whether decreasing plasma TGs without a concomitant decrease in apoB could reduce ASCVD risk by alternative mechanisms unrelated to apoB.

Regarding non-ASCVDs, we show novel causal associations between plasma TG levels and uterine leiomyomas (uterine fibroids), diverticular disease of intestine, paroxysmal tachycardia, hemorrhage from respiratory passages (hemoptysis), and calculus of kidney and ureter (kidney stones). Prior studies had reported correlational associations between many of these diseases and plasma TG levels or risk factors correlated to plasma TGs. For example, studies had documented positive correlations between leiomyoma risk and plasma TG levels (Uimari et al., 2016) (Tonoyan et al., 2021) (Peshkova et al., 2020). Others had shown diverticular disease risk is positively associated with BMI, body fat percentage, and visceral fat area (Shih et al., 2022) (Freckelton et al., 2018) (Böhm, 2021). Similarly, kidney stones have been associated with the triglyceride-glucose (TyG) index (Qin et al., 2021), and atrial tachycardias have been associated with VLDL levels in metabolic syndrome patients (Lee et al., 2017) (Park & Lee, 2018). Our study is the first to suggest that these associations may be causally related and specific to plasma TG levels, as opposed to or in addition to confounding risk factors, such as obesity and hormone replacement therapy. We thus present rationale for repurposing TG-lowering agents towards these diseases and pursuing mechanistic studies interrogating TG biology; however, RCTs are necessary to better evaluate clinical potential.

Our results also identified a novel, negative causal association between plasma TG levels and alcoholic liver disease (ALD), suggesting that excessively reducing plasma TG levels may increase risk of this disease trait. A mechanistic explanation for this association remains elusive as previous studies on dysregulated lipid metabolism in liver disease have generally focused on non-alcoholic fatty liver disease (NAFLD). However, one animal study suggests that medium-chain TGs may decrease lipid peroxidation and reverse established alcoholic liver injury in rat models of ALD (Nanji et al., 1996). Nevertheless, that elevated TG levels may protect against ALD is surprising and requires validation. It remains unclear whether this association is only relevant to lifelong, chronic lowering of plasma TG levels rather than transient lowering by drug-based interventions. It is also unclear whether ALD could be prevented by increasing TG levels. Our finding emphasizes the potential intolerability of TG-lowering agents and the importance of maintaining these drugs’ concentrations within therapeutic windows for patient safety.

This study has several limitations. First, horizontal pleiotropy has been shown to be common in genetic variation, which can cause bias in MR testing (Verbanck et al., 2018) (Jordan et al., 2019). We used MR tests that are robust to horizontal pleiotropy including the MR-PRESSO test as well as MR-Egger and weighted median estimators as sensitivity analyses. Second, genetic variants confer exposures that are lifelong but small in effect size; thus, MR may over- or underestimate the effect sizes of pharmacological interventions. MR also cannot make comparisons between TG-lowering agents as it evaluates drug targets, not drug subclasses and unique pharmacodynamics (Gill et al., 2019) (Sofat et al., 2010). However, the utility of MR in drug target validation is well-established as estimates still indicate the presence and direction of causality (Gill et al., 2021). Third, we were unable to reliably map “categorical” traits from UKB to corresponding traits in FinnGen for all traits, testing for replication only 221 of the 598 associations that were nominally significant in the primary analysis. Moreover, ICD-10 codes and phecodes are imperfect descriptors of disease, liable to misclassification. Lastly, we acknowledge that this study was restricted to populations of European ancestry, limiting the generalizability of our findings. A recent study observed comparable MR estimates for the causal effects of lipid traits on ischemic stroke risk between African and European ancestry individuals (Fatumo et al., 2021), suggesting the potential generalizability of MR results across ancestries in some specific cases. Nevertheless, trans-ancestry MR analyses are warranted to validate our study’s findings in diverse ancestry populations.

In conclusion, this study demonstrates a high-throughput application of two-sample MR to estimate the causal effects of plasma TGs on disease risk phenome-wide. With the proliferation of multiomic data, this systematic approach could be generalized to study the causal effects of serum biomarkers at scale to support the prioritization of targets for drug discovery. Further studies are needed however to consider the functional and qualitative attributes of TRL and TG subtypes. Metabolomic data partitioning TRLs by size and composition may enable this to identify the subcomponent of a plasma TG measurement that drives disease risk (Holmes et al., 2015). Diverse study designs are warranted to further triangulate evidence for the causal effects of plasma TG levels on disease risk.

## Data Availability

All analyses used publicly available data (UKB, FinnGen), including previously published GWAS (GLGC) (Willer et al., 2013). Obtaining access to UKB (Pan-UKB_Team, 2020) and FinnGen (FinnGen, 2020) GWAS summary statistics is detailed here (https://www.finngen.fi/en/access_results) and here (https://pan.ukbb.broadinstitute.org/downloads). Please note the summary statistics for FinnGen and Pan-UKB are made publicly available.

## Acknowledgements

We acknowledge the participants and investigators of the FinnGen and UKB studies.

## Competing interests

RD reports receiving grants from AstraZeneca; grants and non-financial support from Goldfinch Bio; being a scientific co-founder, consultant, and equity holder (pending) for Pensieve Health; and a consultant for Variant Bio, all unrelated to this work. RSR reports receiving grants from Amgen, Arrowhead, Lilly, Novartis and Regeneron; consulting fees from Amgen, Arrowhead, Lilly, Novartis and Regeneron; honoraria for non-promotional lectures from Amgen, Kowa and Regeneron, royalties from Wolters Kluwer (UpToDate); and stock holdings in MediMergent, LLC.

## Supplementary files

**S1 Table. Causal estimates of plasma TG levels on 2,600 traits in UKB using multiple MR methods.**

Shown are the estimates, standard deviations, and *p*-values of MR results using IVW, MR-Egger, and Weighted Median methods. 141 SNPs were used as instrumental variables to proxy plasma TG levels. A Bonferroni threshold for statistical significance was set to *p* < 0.05 / 2600 = 1.92 × 10^-5^ for this discovery analysis.

**S2 Table. Causal estimates of plasma TG levels on 221 traits in FinnGen using multiple MR methods.**

Shown are the estimates, standard deviations, and *p*-values of MR results using IVW, MR-Egger, and Weighted Median methods. 1,248 SNPs were used as instrumental variables to proxy plasma TG levels. A Bonferroni threshold for statistical significance was set to *p* < 0.05 / 221 = 2.26 × 10^-4^ for this replication analysis.

**S3 Table. IVW-MR estimates of significant and replicated associations (Tier 1-3) after MR-PRESSO outlier tests using UKB data.**

Shown are the estimates, standard deviations, and *p*-values of IVW-MR results in the discovery analysis, before and after outlier IV removal, for significant and replicated traits (tier 1-3) that had significant MR-PRESSO global test results (*p* < 0.05) in the primary IVW analysis of the discovery UKB cohort.

**S4 Table. Multivariable IVW-MR estimates of plasma TG levels on significant and replicated associations (Tier 1-3) using UKB data.**

Shown are the estimates, standard deviations, and *p*-values of multivariable IVW-MR results controlling for HDL-C, LDL-C, or both, in the discovery stage. Only tier 1-3 results significant and replicated as predefined in the methods were examined in this analysis.

## Notes

Funding: RD is supported by the National Institute of General Medical Sciences of the National Institutes of Health (NIH) (R35-GM124836) and the National Heart, Lung, and Blood Institute of the NIH (R01-HL139865 and R01-HL155915).

### Funding Statement

RD is supported by the National Institute of General Medical Sciences of the National Institutes of Health (NIH) (R35-GM124836) and the National Heart, Lung, and Blood Institute of the NIH (R01-HL139865 and R01-HL155915).

